# Exclusive Breastfeeding at 6 Weeks Related to Pain Self-Management, Emotion and Self-Efficacy: A Secondary Analysis of a Pilot Randomized Control Trial

**DOI:** 10.1101/2020.09.28.20195198

**Authors:** Ruth Lucas, Yiming Zhang, Beth Russell, Pornpan Srisopa, Julianna Boyle, Jimi Francis

**Affiliations:** School of Nursing, University of Connecticut; Department of Statistics, University of Connecticut; Human Development and Family Science, University of Connecticut; Department of Health & Kinesiology, University of Texas at Tyler

**Keywords:** Exclusive breastfeeding, women, pain self-management, pain self-management intervention, emotion

## Abstract

**Introduction:** Exclusive breastfeeding (EBF) outcomes can vary by concomitant emotions, ongoing pain, and breastfeeding self-efficacy. The purpose of this study is to examine the association of ongoing pain with breastfeeding, concomitant emotions and breastfeeding self-efficacy scores (BSES) with EBF outcomes at 6 weeks postpartum.

**Design:** A secondary analysis of a randomized pilot trial of a home-based breastfeeding pain self-management (BSM) intervention for 56 mothers (26 BSM, 30 Control). The BSM intervention provided self-management strategies for breastfeeding and breastfeeding pain. Effect modification of EBF and associated symptoms of depression, anxiety, sleep, well-being pain severity scores, BSES, and group assignment was assessed using the likelihood ratio test.

**Results:** EBF at 6 weeks controlling for demographic covariates, the group, pain severity, anxiety and sleep were significant predictors. Mothers with every one point increase in their pain severity score and sleep score, had a decrease of 9% (OR = .91, 95%CI = [.79, .98]) and 25% (OR = .75, 95% CI = [.52, .95]) respectively and with a one point increase in the anxiety score, a 58% (OR = 1.58, 95% CI = [1.13, 2.72]) increase in the odds of EBF at week 6.

**Conclusion:** Examinations of EBF at 6 weeks should include evaluation of mothers’ ongoing pain and emotional distress, as mothers continue breastfeeding even at personal cost. Early identification and validation of breastfeeding challenges, ongoing pain, and emotional distress are needed to bolster mothers’ confidence in their breastfeeding skills, thus supporting their EBF goals.

**Quick points:**

- Unresolved pain during breastfeeding may affect breastfeeding self-efficacy, maternal self-efficacy and bonding
- The effect of maternal emotion on breast and nipple pain and breastfeeding outcomes is not consistently evaluated during breastfeeding
- Mothers who received breastfeeding pain self-management strategies reported higher breastfeeding self-efficacy scores and lower pain and anxiety scores
- Providing breastfeeding pain self-management is a pathway to increase breastfeeding self-efficacy and exclusive breastfeeding
- Empowering women with the skills needed to self-manage anxiety and pain by providing real-time support in the first weeks after birth support mother to reach their breastfeeding goals

---

For some mothers, the “natural” task of breastfeeding can be a roller coaster of emotions and may include physical aspects such as tenderness of breast and nipple tissue. Although tenderness might be expected for a short time as lactating tissue transitions from inactivity to human milk production and breastfeeding, pain should not be a part of breastfeeding. Pain during lactation is an indicator of malfunction and is a leading cause of distress for more than 1 million mothers, heavily influencing feeding decisions and early weaning.^1,2^ Additional studies have confirmed that mothers who continue to experience pain as lactation progresses are at greater risk for increased mood disturbances, increased interference with activities of daily living, and changes in sleep cycle.^3,4^ Unresolved pain during breastfeeding may also affect breastfeeding self-efficacy, maternal self-efficacy and bonding which may have an enduring and negative impact on the maternal/infant dyad.^4–6^

Emotional reactions to pain can suppress or amplify the perception of painful stimuli. In fact, emotion is included in the 1994 definition of pain adopted by the International Association for the Study of Pain. Over the last 25 years, significant associations between pain and negative emotion have become well established.^7,8^ The connections between emotional reactions and painful stimuli are often described through approach-avoidance models when achieving a goal involving both positive and negative experiences. Employing these models in an intervention design can reduce the obstacles psychological stress, depression, and anxiety often seen in chronic disease management^9^, and to new mothers’ abilities to manage their health and comfort while pursuing their quality of life goals to breastfeed their infant over the weeks and months after birth.^5^

Uniquely taxing for mothers’ emotional regulation skills are the dysregulating effects of sleep/wake cycle disruptions.^10,11^ Breast and nipple pain is associated with changes in sleep cycles as well as increased pain interference with activities of daily living and maternal mood.^12^ The strain of managing breastfeeding in this sleepless context can cause distress and interfere with mothers’ ability to 1) focus on her postpartum needs, 2) impair problem-solving, 3) prioritize, and finally 4) engage in self-management activities.^13^ Relatedly, breastfeeding self-efficacy reflects maternal confidence to manage breastfeeding and meet their breastfeeding goals. Dennis (1999) theorized that breastfeeding self-efficacy includes the confidence to successfully manage pain and emotions, particularly anxiety during breastfeeding.^14^ Yet, it is clear that with increased pain, there is decreased breastfeeding self-efficacy, increased emotional distress, and a higher rate of mothers not reaching their breastfeeding goals.^2,12^

## Current Study & Research Aims

Our pilot randomized trial (RCT) used a breastfeeding pain self-management (BSM) intervention to target pain self-management and to support EBF.^15^ The goal of the study was to decrease mothers’ breast and nipple pain using self-management strategies. The BSM intervention significantly decreased breastfeeding pain during the first two weeks postpartum.^16^ However, at 6 weeks, there was no significant difference between groups for ongoing pain (*p* = .368) and EBF (*p*=.092). This secondary analysis explores the associations between EBF and concomitant symptoms of depression, anxiety, sleep, general well-being, and breastfeeding self-efficacy at 6 weeks as moderated by the BSM intervention. We hypothesize that EBF at 6 weeks will be associated with decreased pain and symptoms of depression, anxiety and sleep disruption and increased self-efficacy and perceived well-being.

## Methods

### Study Design

This report is a secondary analysis of selected data collected from the pilot RCT BSM intervention among breastfeeding mothers in 2017. All study materials and protocol were approved by the University of Connecticut Institutional Review Board and registered with Clinical Trials.gov (NCT03392675).

### Setting

Participants were recruited at two research-intensive regional tertiary medical centers in the northeast region of the United States.

### Sample

A convenience sample of 78 participants were approached and 65 participants consented during recruitment before hospital discharge and followed-up at 1, 2, and 6 weeks after delivery. All study personnel involved in data collection remained blinded to the group assignment of participants. Five mothers (BSM intervention) consented but did not complete the initial documents precluding enrollment, and four mothers (1 BSM, 3 Control) stopped breastfeeding before 6 weeks. A total of 56 mothers (26 BSM, 30 Control via computer-generated randomization scheme generated by the study statistician) completed surveys and were breastfeeding at 6 weeks (Figure 1). These were included in this analysis. The pilot RCT was a feasibility study, with a sample size goal of 60 mothers which was large enough to report significant differences in average breastfeeding pain severity scores between the BSM and Control groups ^17^.

Inclusion criteria for this study mothers 1) 18 - 45 years of age; 2) breastfeeding to at least 6 weeks; 3) with a full-term infant (38 – 42 gestational weeks) without medical complications; 4) read and speak English, and 5) had daily access to a smartphone or computer. Exclusion criteria included 1) mothers who had a history of potential changes in pain sensorium such as significant mental health disorder (i.e. schizophrenia, bipolar disorder); 2) health condition(s) not associated with pregnancy (i.e. sickle cell anemia, HIV+, diabetes, history of seizures); 3) delivered twin infants, or an infant with congenital anomalies or ankyloglossia that would interfere with breastfeeding; and 4) stopped breastfeeding before 6 weeks.

### Measurements

Participants completed assessments of breast and nipple pain intensity using a horizontal visual analogue scale with fixed intervals between 0-100, and reported frequency and type of daily feedings (breast, human milk in a bottle, or formula) at baseline and 6 weeks.^15^

The 14-item Breastfeeding Self-Efficacy Scale - Short Form (BSES-SF) assessed maternal confidence with breastfeeding was completed at baseline and 6 weeks. BSES scores > 50 indicate greater maternal breastfeeding confidence (α =.967).^18^

Maternal emotion was assessed by the 6-item Patient Reported Outcomes Measurement Information System (PROMIS®) for anxiety (α =.824), and sleep (α = .819) was measured at baseline and 6 weeks (*PROMIS Instrument*, n.d.). The 10-item PROMIS Global Health with Cronbach’s α = .584, was measured overall wellbeing (quality of life, mental health, satisfaction with social activities, and emotional problems) at baseline and 6 weeks.^20^ Depressive symptoms were measured by the Edinburgh Postnatal Depression Scale (EPDS) at baseline and 6 weeks. The EPDS is a 10-item scale with each item having 4 choices (0 – 4). Any total score >10 indicates risk for possible depression and scores > 13 a depressive illness that requires referral. The scale has a sensitivity of 85%, with a specificity of 77% to identify mothers high-risk for depression and for our study, a Cronbach’s α = .813 ^21^.

### Data Collection

The BSM intervention was implemented from hospital discharge to 6 weeks from April 2017 to October 2017, and included bi-weekly nurse-led texting, access to online educational modules targeting knowledge and beliefs to self-manage breastfeeding and breast and nipple pain, and a breastfeeding journal. Mothers in the intervention group also received bi-weekly cognitive therapy-based educational modules addressing challenges in breastfeeding and examples as to how to manage breast and nipple discomfort, and hyperlinks to online resources for the first 2 weeks from discharge. The Control group had access to usual care, including access to an outpatient lactation consultant. Both the BSM intervention and the Control group received text/email at 1, 2, and 6 weeks, with a link to complete assessments for maternal report of breast and nipple pain severity, symptoms of anxiety, depression, and sleep disruption, breastfeeding self-efficacy, and breast and formula feeding frequency. Additionally, the PROMIS measures for Anxiety, Sleep, and Global Health were completed at baseline and 6 weeks.

### Data Analysis

Independent two sample t-tests on continuous variables and Pearson Chi-Square test or Fisher’s exact test on discrete variables were performed to verify the non-significant difference of demographic characteristics between the two groups. Key variables of interest were assessed to determine whether the assumptions of normality were met. We conducted the paired two sample t-tests to examine changes in the key variables over time. Logistic regression was used to test study hypotheses since the EBF is a dichotomous outcome. We put group, depression, pain severity, anxiety, sleep, and BSES at 6 weeks as the explanatory variables, with controlling age, ethnicity, education, marital status, employment and if breastfed before as adjusting covariates. Likelihood ratio test (LRT) was conducted to detect the significance of each explanatory variable in the logistic regression model.

## Results

In comparing the two groups at baseline, there were no significant differences in demographic data (see Table 1). In addition, the paired two sample t-tests of key variables indicated significant improvements of BSES, well-being (Global Health), Sleep, Anxiety and pain severity within BSM and Control groups and as a total group from baseline to 6 weeks (see Table 2). Further, there were no group differences for Global Health, Pain Severity, Anxiety, BSES or EPDS scores at either baseline or 6 weeks.

**Table 1.**
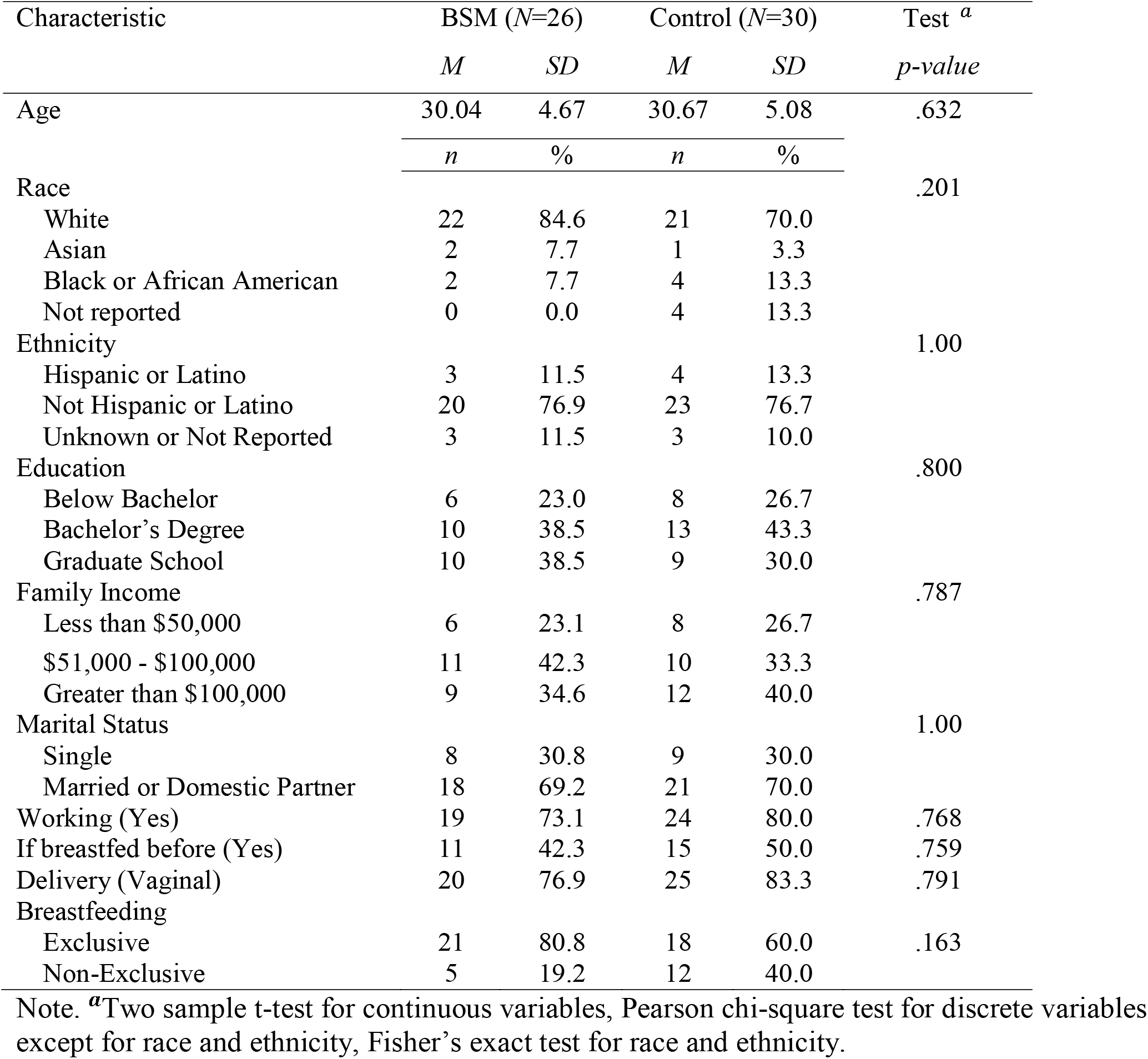
Demographic Characteristics by Group.

**Table 2.**
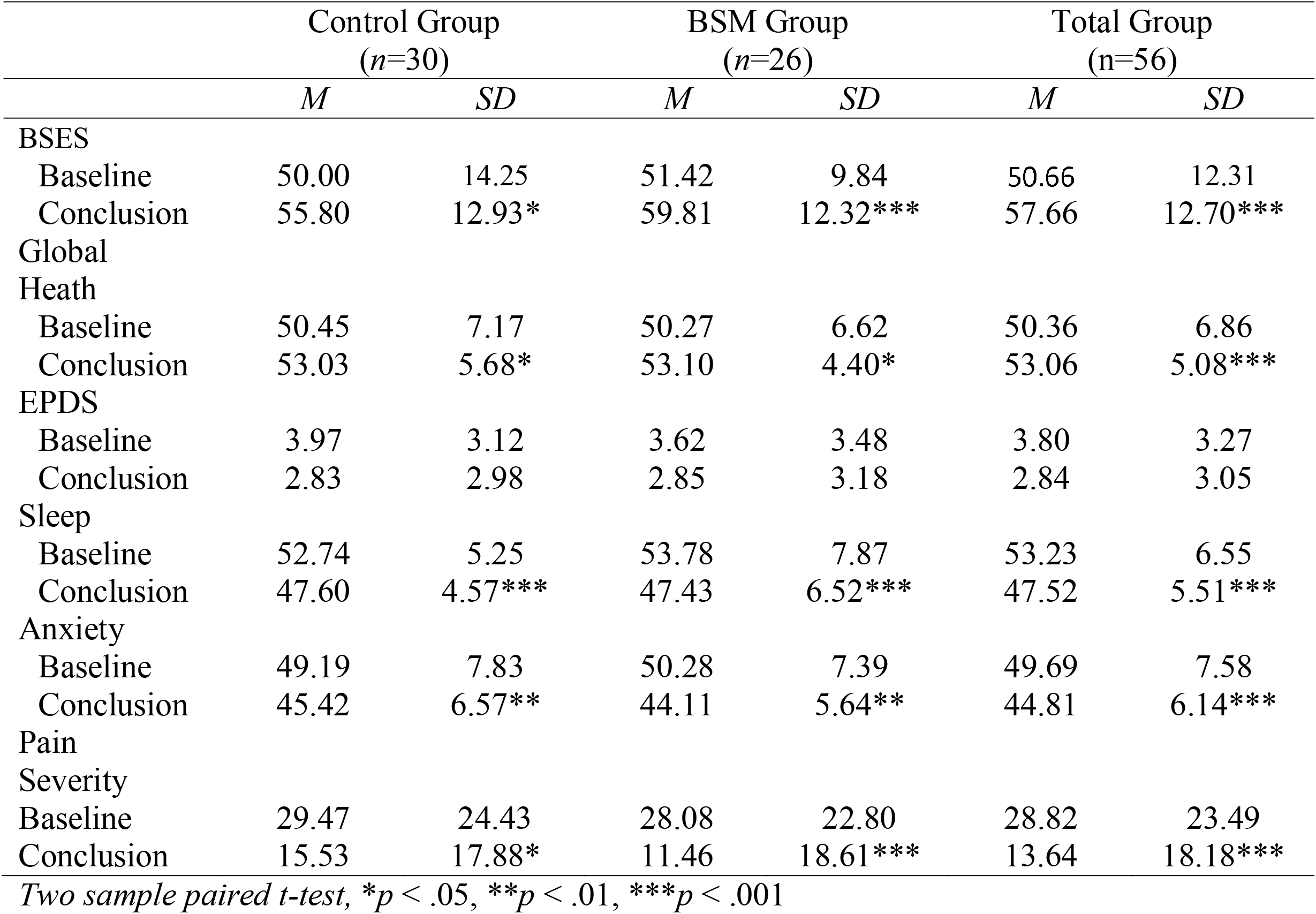
Key Variables - Score Changes from Baseline to 6 Weeks by Group (N=56)

|  | Control Group<br>(n=30) |  | BSM Group<br>(n=26) |  | Total Group<br>(n=56) |  |
| --- | --- | --- | --- | --- | --- | --- |
|  | *M* | *SD* | *M* | *SD* | *M* | *SD* |
| BSES |  |  |  |  |  |  |
| Baseline | 50.00 | 14.25 | 51.42 | 9.84 | 50.66 | 12.31 |
| Conclusion | 55.80 | 12.93* | 59.81 | 12.32*** | 57.66 | 12.70*** |
| Global |  |  |  |  |  |  |
| Heath |  |  |  |  |  |  |
| Baseline | 50.45 | 7.17 | 50.27 | 6.62 | 50.36 | 6.86 |
| Conclusion | 53.03 | 5.68* | 53.10 | 4.40* | 53.06 | 5.08*** |
| EPDS |  |  |  |  |  |  |
| Baseline | 3.97 | 3.12 | 3.62 | 3.48 | 3.80 | 3.27 |
| Conclusion | 2.83 | 2.98 | 2.85 | 3.18 | 2.84 | 3.05 |
| Sleep |  |  |  |  |  |  |
| Baseline | 52.74 | 5.25 | 53.78 | 7.87 | 53.23 | 6.55 |
| Conclusion | 47.60 | 4.57*** | 47.43 | 6.52*** | 47.52 | 5.51*** |
| Anxiety |  |  |  |  |  |  |
| Baseline | 49.19 | 7.83 | 50.28 | 7.39 | 49.69 | 7.58 |
| Conclusion | 45.42 | 6.57** | 44.11 | 5.64** | 44.81 | 6.14*** |
| Pain |  |  |  |  |  |  |
| Severity |  |  |  |  |  |  |
| Baseline | 29.47 | 24.43 | 28.08 | 22.80 | 28.82 | 23.49 |
| Conclusion | 15.53 | 17.88* | 11.46 | 18.61*** | 13.64 | 18.18*** |

*Two sample paired t-test, \*p < .05, \*\*p < .01, \*\*\*p < .001*

Table 3 presents the estimation of coefficients for the logistic regression which modelled the EBF at 6 weeks with Group, BSES, pain severity, anxiety, depression, sleep and wellbeing (Global Health). With adjusting for the important demographic covariates including age, ethnicity, education, marital status, employment and if breastfed before, the group (B = 2.42, p = .043), pain severity (B = -.10, p = .014), anxiety (B = .45, p = .003) and sleep (B = -.29, p = .016) were significant predictors in the model (Table 3). Holding all other covariates in the model fixed, for a one point increase in pain severity score and sleep score, we expect mothers to have 9% (OR = .91, 95%CI = [.79, .98]) and 25% (OR = .75, 95% CI = [.52, .95]) decrease respectively in the odds of EBF at week 6. For a one point increase in the anxiety score, we expect mothers to have 58% (OR = 1.58, 95% CI = [1.13, 2.72]) increase in the odds of EBF at week 6.

**Table 3.** Concomitant Emotions and Wellbeing Associated with EBF (N = 56)

| Variable | *B* | OR | 95% CI of OR |
| --- | --- | --- | --- |
| Group | 2.42 | 11.28 | [1.08, 27.43] |
| BSES | .09 | 1.09 | [1.00, 1.22] |
| Pain severity | -.10 | .91 | [.79, .98] |
| Anxiety | .45 | 1.58 | [1.13, 2.72] |
| EPDS | -.17 | .84 | [.48, 1.40] |
| Sleep | -.29 | .75 | [.52, .95] |
| Global Health | -.18 | .84 | [.62, 1.08] |

Note: OR = Odds Ratio. CI = Confidence Interval. EBF = Exclusive Breastfeeding. Logistic regression controlled for age, ethnicity, education, marital status, employment and if breastfed before.
<sup>a</sup> Likelihood Ratio Test (LRT) p-value.

## Discussion

Mothers in the BSM intervention group had higher BSES scores and lower pain and anxiety scores at 6 weeks compared to the Control group. Mothers in the intervention group received early validation that their breastfeeding pain was real and were provided with self-management strategies to address their pain and anxiety during breastfeeding across the 6 weeks. In contrast, mothers in the Control group required 6 weeks to decrease their pain severity scores and reported lower BSES and higher anxiety scores. These results support the hypothesis that EBF was associated with group, decreased pain, but contrary to the literature, increased anxiety, which represents an opportunity to address the self-management of breastfeeding pain as a pathway to managing anxiety and increasing breastfeeding self-efficacy ^22–24^.

At 6 weeks, 36% of participants did not have improvement or resolution of their pain. Taken together, these findings indicate mothers will persist with breastfeeding even at great personal cost. Based on bi-variate results, we found that mothers in the intervention group were benefited by managing their pain and sleep symptoms and were significantly more likely to be EBF at 6 weeks. Our findings support Dennis’ theory (1999) that increased breastfeeding self-efficacy is an outcome of the self-management of pain, however increased, not decreased, anxiety was a significant contribution to EBF at 6 weeks. The presence of anxiety at 6 weeks may be multifactorial as mothers are preparing to return to work, often pumping additional breast milk to be stored for future feedings, and for a third of the mothers, continuing to experience pain. Given the addition of wellbeing did not strengthen the prediction of breastfeeding outcomes, our results underscore the powerful impact of negative emotion experiences in the postpartum period. In this light, a woman’s ability to regulate her emotions is pivotal to breastfeeding success and to her recollection of her breastfeeding goals and informing her future breastfeeding decisions ^1,23^.

Russell and Park (20^18^) describe the ability to self-manage positive and negative emotions as pivotal to resilience during intense, transient, or continuously stressful life events. In comparison to other clinical pain conditions, postpartum mothers also experience ongoing breastfeeding pain and concomitant symptoms of anxiety, fatigue, and depression; these may affect pain perception and decrease health self-efficacy ^14,22,25^. Breastfeeding is a unique health behavior due to its cultural value, powerful health impacts, and because it has become common in the United States for a proportion of mothers to experience extended pain and discomfort. Mothers understand that breastfeeding is pivotal for the infant’s health, maternal-infant attachment, and is culturally important ^23^. In our larger study, although mothers experienced pain during breastfeeding at 1 and 2 weeks, there was no significant relationship between early discomfort and exclusive breastfeeding at 6 weeks ^15^. This result suggests that mothers may continue breastfeeding despite experiencing pain and discomfort that endures for multiple weeks. This result is in line with Schwartz et al. (2002), who found that each day of pain during the first 3 weeks of breastfeeding increased the risk of discontinuing breastfeeding by 10 – 26% (leading to formula use and lower maternal self-efficacy), however, after 3 weeks, mothers no longer reported pain as a reason for breastfeeding cessation. In fact, mothers who persevered through pain, express feelings of empowerment, but may still experience adverse emotions, such as anxiety ^26^. However, for some women with unresolved emotions may have an enduring and negative impact on the maternal/infant dyad ^4,6^.

The consequences of breast and nipple pain extend far beyond physical and neurosensory systems and intrude into the whole of a woman’s life with impacts on her affective, cognitive, and social functioning ^3–5^. Thus, breast and nipple pain interventions need to integrate emotion regulation considerations in self-management plans for breast and nipple pain in order to increase mothers’ ability to modulate or persevere in the face of pain ^1,5,23^. Mothers’ negative emotional experiences do affect their personal breastfeeding perception and breastfeeding outcomes. In several studies, mothers’ anxiety and depression after delivery was associated with earlier introduction of formula and breastfeeding cessation before 3 months ^5,27^. Thus the management of maternal anxiety is pivotal to EBF beyond 6 weeks ^24^.

These findings reinforce the importance of providing real-time support for mothers in the first weeks after birth to realize their breastfeeding goals against any pain, discomfort, and resulting emotional distress. Specifically, when mothers are psychologically distressed (experiencing stress, depression, or anxiety), they experience increased illness symptomology including pain ^28^, and may form inaccurate illness perceptions that interfere with adaptive self-management behaviors ^22,23^. Birth and establishing breastfeeding require resilience and self-management, yet the effect of psychological distress is often not considered as a contributor to continuing breast and nipple discomfort and pain ^16,29^. Further, there is evidence that psychological distress is the single best predictor of pain-related disability ^29^.

Our results suggest there may be a set of teachable self-management skills that help breastfeeding mothers persist despite enduring painful experiences. At week 6, EBF were significantly associated improved symptoms of pain severity anxiety, and sleep disturbances, aligning with the literature ^16,24,30^. These findings are also aligned with qualitative evidence from Kronborg and colleagues (2015) who identify key themes in mothers’ narrative themes in establishing a breastfeeding-bonding trajectory that highlight anxious breastfeeding experiences including insecurity, worry, and fear (i.e., concerns about human milk production, social approval, and feeding capability). Future studies might consider intervening with targeted emotional support in addition to breastfeeding self-management skills during the first 1 to 2 weeks and continue until emotional support until at least 6 weeks. For clinicians, it is important to validate mothers’ report of breast and nipple pain and provide mothers with reassurance regarding their anxiety related to breastfeeding. Together, these will decrease ongoing breastfeeding pain and support mothers’ realization of their breastfeeding goals.

### Limitations

Our study findings are limited due to sample size and the secondary analysis design. We did not assess emotion regulation constructs each week, including anxiety as a key variable of interest. Further, at 6 weeks issues related to breastfeeding were resolving. Our pilot findings strongly suggest that the pivotal time for addressing pain self-management and its emotional correlates in an intervention is in the first weeks directly following birth when pain is a significant risk for stopping breastfeeding ^5,16,23^.

## Conclusion

The consequences of breast and nipple pain extend far beyond physical and neurosensory systems and intrude into the whole of a mother’s life, in affective, cognitive, and social functions. Breast and nipple pain interventions need to integrate self-management for specific conditions related to breast and nipple pain with emotion regulation in order to increase mothers’ ability to regulate and modify pain in all dimensions. Empowering mothers with the skills needed to self-manage anxiety and pain by providing real-time support in the first weeks after birth could lead to mothers to improved realization of their breastfeeding goals. Mothers seeking support for pain need to have the validation from their care providers that their pain is real but a manageable challenge. It is imperative that resources are available to assist mothers to set expectations and provide support should pain and its associated emotional experiences persist. Additional work needs to be done to find effective ways to collaborate with mothers about their expectations and how they manage their pain if it develops and persists. This work is crucial to developing self-management interventions that acknowledge and consider emotional experiences that may arise when they persist even when “it hurts”.

## Data Availability

Deidentified symptom data (including the data dictionary) will be provided to interested scientific investigators for the purpose of secondary research analysis by contacting the corresponding author and sending the investigators curriculum vitae and research questions. The study protocol and statistical analysis plan is available in our previous publication.

https://cdrns.nih.gov/how-to/access-data

## CONSORT 2010 checklist of information to include when reporting a randomised trial*

CONSORT 2010 checklist of information to include when reporting a randomised trial\*
| Section/Topic | Item No | Checklist item | Reported on page No |
| --- | --- | --- | --- |
| **Title and abstract** |  |  |  |
|  | 1a | Identification as a randomised trial in the title | 1 |
|  | 1b | Structured summary of trial design, methods, results, and conclusions (for specific guidance see CONSORT for abstracts) | 2 |
| **Introduction** |  |  |  |
| Background and objectives | 2a | Scientific background and explanation of rationale | 3 |
|  | 2b | Specific objectives or hypotheses | 4 |
| **Methods** |  |  |  |
| Trial design | 3a | Description of trial design (such as parallel, factorial) including allocation ratio | 5 |
|  | 3b | Important changes to methods after trial commencement (such as eligibility criteria), with reasons | 5 |
| Participants | 4a | Eligibility criteria for participants | 5 |
|  | 4b | Settings and locations where the data were collected | 5 |
| Interventions | 5 | The interventions for each group with sufficient details to allow replication, including how and when they were actually administered | 6 |
| Outcomes | 6a | Completely defined pre-specified primary and secondary outcome measures, including how and when they were assessed | 4 |
|  | 6b | Any changes to trial outcomes after the trial commenced, with reasons | - |
| Sample size | 7a | How sample size was determined | 5 |
|  | 7b | When applicable, explanation of any interim analyses and stopping guidelines | - |
| Randomisation: |  |  |  |
| Sequence generation | 8a | Method used to generate the random allocation sequence | 5 |
|  | 8b | Type of randomisation; details of any restriction (such as blocking and block size) | - |
| Allocation concealment mechanism | 9 | Mechanism used to implement the random allocation sequence (such as sequentially numbered containers), describing any steps taken to conceal the sequence until interventions were assigned | - |
| Implementation | 10 | Who generated the random allocation sequence, who enrolled participants, and who assigned participants to interventions | 5 |
| Blinding | 11a | If done, who was blinded after assignment to interventions (for example, participants, care providers, those assessing outcomes) and how | 5 |
|  | 11b | If relevant, description of the similarity of interventions | - |
| Statistical methods | 12a | Statistical methods used to compare groups for primary and secondary outcomes | 7 |
|  | 12b | Methods for additional analyses, such as subgroup analyses and adjusted analyses | y |
| **Results** |  |  |  |
| Participant flow (a diagram is strongly recommended) | 13a | For each group, the numbers of participants who were randomly assigned, received intended treatment, and were analysed for the primary outcome | 8 |
|  | 13b | For each group, losses and exclusions after randomisation, together with reasons | Cohort |
| Recruitment | 14a | Dates defining the periods of recruitment and follow-up | 7 |
|  | 14b | Why the trial ended or was stopped | 7 |
| Baseline data | 15 | A table showing baseline demographic and clinical characteristics for each group | Table 1 |
| Numbers analysed | 16 | For each group, number of participants (denominator) included in each analysis and whether the analysis was by original assigned groups | 5 |
| Outcomes and estimation | 17a | For each primary and secondary outcome, results for each group, and the estimated effect size and its precision (such as 95% confidence interval) | - |
|  | 17b | For binary outcomes, presentation of both absolute and relative effect sizes is recommended | - |
| Ancillary analyses | 18 | Results of any other analyses performed, including subgroup analyses and adjusted analyses, distinguishing pre-specified from exploratory | - |
| Harms | 19 | All important harms or unintended effects in each group (for specific guidance see CONSORT for harms) | - |
| **Discussion** |  |  |  |
| Limitations | 20 | Trial limitations, addressing sources of potential bias, imprecision, and, if relevant, multiplicity of analyses | 12 |
| Generalisability | 21 | Generalisability (external validity, applicability) of the trial findings |  |
| Interpretation | 22 | Interpretation consistent with results, balancing benefits and harms, and considering other relevant evidence | 12 |
| **Other information** |  |  |  |
| Registration | 23 | Registration number and name of trial registry | 1 |
| Protocol | 24 | Where the full trial protocol can be accessed, if available | 1 |
| Funding | 25 | Sources of funding and other support (such as supply of drugs), role of funders | 1 |

\*We strongly recommend reading this statement in conjunction with the CONSORT 2010 Explanation and Elaboration for important clarifications on all the items. If relevant, we also recommend reading CONSORT extensions for cluster randomised trials, non-inferiority and equivalence trials, non-pharmacological treatments, herbal interventions, and pragmatic trials. Additional extensions are forthcoming: for those and for up to date references relevant to this checklist, see www.consort-statement.org.

## CONSORT 2010 Flow Diagram

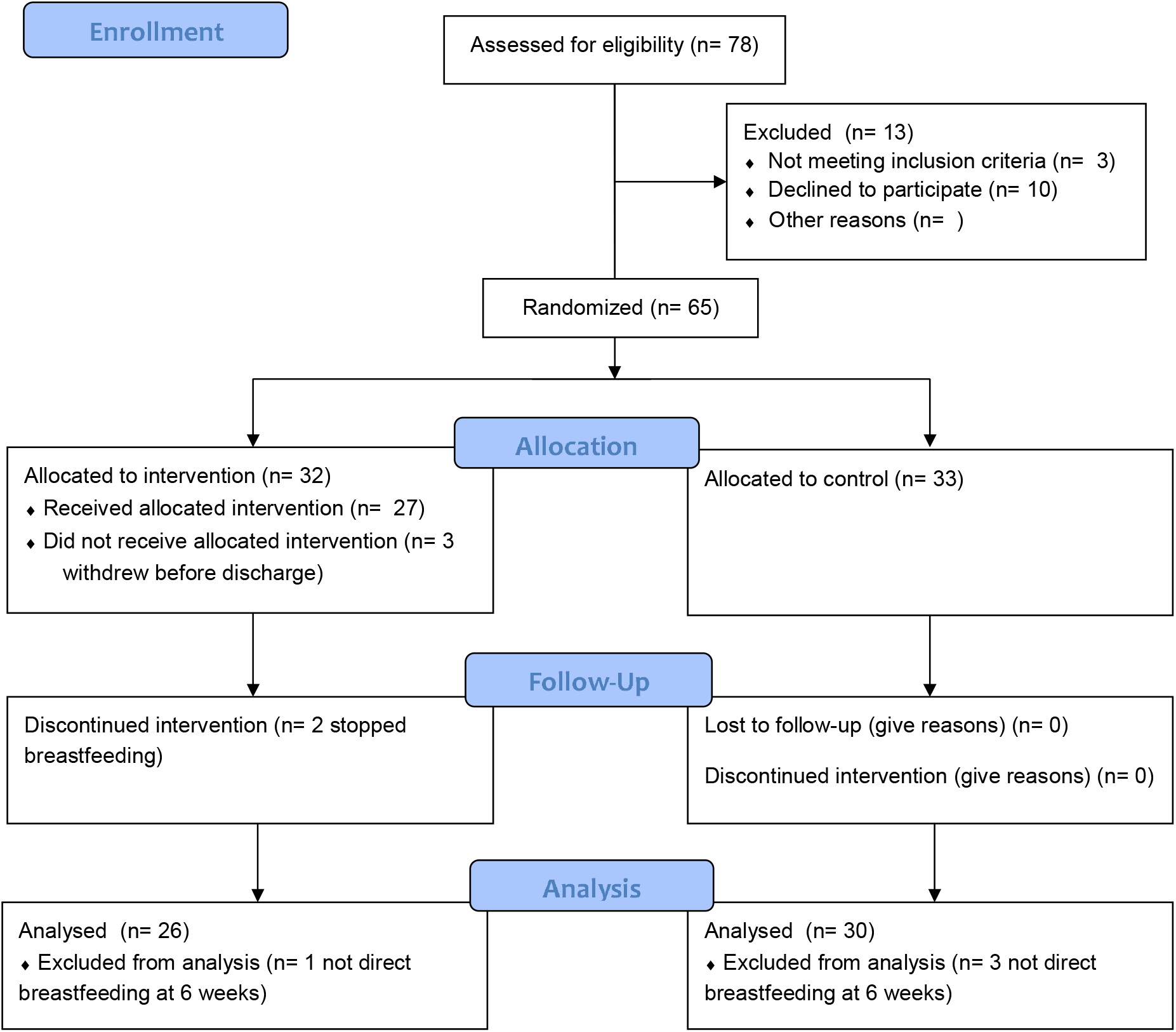

## Notes

Supported by the National Institutes of Health-NINR P20NR016605 to Dr. Angela Starkweather as primary investigator and Ruth Lucas as pilot primary investigator. Clinical trial conducted at the University Of Connecticut School Of Nursing. Jimi Francis and Ruth Lucas are co-owners of Dyadic Innovations, LLC. They have no conflicts of interest to disclose related to Dyadic Innovations, LLC. Yiming Zhang, Beth Russell, Julianna Boyle, and Pornpan Srisopa have not conflicts of interest to disclose. Deidentified symptom data (including the data dictionary) will be provided to interested scientific investigators for the purpose of secondary research analysis by contacting the corresponding author and sending the investigator’s curriculum vitae and research questions. The study protocol and statistical analysis plan is available in our previous publication.

### Competing Interest Statement

The authors have declared no competing interest.

### Clinical Trial

NCT03392675

### Clinical Protocols

https://onlinelibrary.wiley.com/doi/abs/10.1002/nur.21938

### Funding Statement

NINR P20NR016605

### Author Declarations

The University of Connecticut Institutional review board (IRB) approved the study (H16-199) prior to enrolling participants.

